# Effect of a culturally adapted birth preparedness and complication readiness training intervention on knowledge among Hehe pregnant women in rural Iringa Tanzania: A controlled pretest–posttest quasi-experimental study

**DOI:** 10.64898/2026.10.13.26365390

**Authors:** Rosalia B. Mwenda, Saada Ali Seif, Rehema O. Stephano, Fabiola V. Moshi

**Affiliations:** Department of Nursing Management and Education, School of Nursing and Public Health, University of Dodoma, Tanzania; Department of Public Health, School of Nursing and Public Health, University of Dodoma, Tanzania; Department of Kiswahili, University of Dodoma, Tanzania; Department of Clinical Nursing, School of Nursing and Public Health, University of Dodoma, Tanzania

**Keywords:** Birth preparedness, Culturally adapted health education, BPCR knowledge, Antenatal care, Pregnant women, Cultural and linguistic adaptation, Quasi-experimental study

## Abstract

**Background:** Birth preparedness and complication readiness (BPCR) knowledge is important for recognizing maternal and newborn danger signs and preparing for timely care. However, conventional translation of BPCR educational materials may not adequately address local linguistic and cultural contexts. This study evaluated whether a culturally adapted Swahili BPCR training manual improved knowledge more than a standard translated Swahili BPCR training manual among Hehe pregnant women in rural Iringa, Tanzania.

**Methods:** A controlled pretest–posttest quasi-experimental study was conducted among 176 pregnant women attending antenatal care in Mufindi and Kilolo districts. Eighty-eight women received the Culturally Adapted Swahili BPCR Training Manual and 88 received the Standard Translated Swahili BPCR Training Manual. Both groups received comparable BPCR education with standardized session duration, delivery format, language, and follow-up procedures. BPCR knowledge was assessed using a 64-item questionnaire at baseline and four weeks after the intervention. The primary analysis used Difference-in-Differences regression to compare changes in knowledge between groups. Multiple linear regression was used to assess the association between intervention-group assignment and post-intervention knowledge after adjustment for baseline knowledge and participant characteristics.

**Results:** All 176 participants completed both assessments. Mean knowledge scores increased from 21.01 (SD 7.42) to 38.49 (SD 7.54) in the culturally adapted group and from 22.51 (SD 5.80) to 32.81 (SD 5.55) in the standard translated group. The culturally adapted group demonstrated a 7.18-point greater increase in knowledge than the standard translated group (β = 7.18, 95% CI: 4.40–9.97; p < 0.001). After adjustment for baseline knowledge, age, education, gravidity, marital status, living arrangement, reading ability, and occupation, intervention-group assignment remained significantly associated with higher post-intervention knowledge (β = 5.88, 95% CI: 3.84–7.92; p < 0.001).

**Conclusion:** Culturally adapted BPCR education was associated with greater short-term improvement in knowledge than standard translated education among Hehe pregnant women. Integrating culturally and linguistically appropriate terminology, contextual explanations, and locally meaningful communication into BPCR education may strengthen knowledge outcomes. Multisite studies with stronger allocation procedures and longer follow-up are needed to confirm these findings.

**Trial Registration:** Pan African Clinical Trials Registry: PACTR202607626025411. Registered retrospectively on 09 July 2026.

## Background

Maternal and neonatal mortality remain major public health challenges, particularly in low- and middle-income countries. Globally, an estimated 260,000 women die from maternal causes each year, with sub-Saharan Africa accounting for approximately 70% of these deaths (1, 2). Tanzania continues to experience a substantial maternal health burden despite improvements in access to antenatal and maternity services. The national maternal mortality ratio was estimated at 104 per 100,000 live births, while 90% of pregnant women were attended by skilled birth attendants and 78% attended at least one antenatal care visit (3, 4). Maternal deaths are commonly associated with delays in recognizing complications, deciding to seek care, reaching health facilities, and receiving adequate treatment (5, 6).

Knowledge of pregnancy, childbirth, postpartum, and newborn danger signs is essential for timely recognition of complications and appropriate care-seeking (7, 8). Birth Preparedness And Complication Readiness (BPCR) is an established approach that promotes recognition of danger signs and advance preparation for skilled birth attendance, transportation, financial resources, and other requirements for emergency care (9–11) However, substantial gaps in BPCR knowledge have been reported in Tanzania and other sub-Saharan African settings. In Iringa, Tanzania, a baseline study found a mean BPCR knowledge score of 21.59 out of 62 (34.8% of the maximum possible score) (9), while only 14% of pregnant women in Mwanza had adequate BPCR knowledge (12). Similar gaps have been reported in Kenya, Ethiopia, and Uganda (13–15).

The effect of maternal health education may depend not only on the provision of information but also on whether it is understood within recipients’ cultural and linguistic contexts (16, 17). Culturally appropriate health communication can incorporate locally meaningful terminology, examples, and explanations to improve comprehension and engagement with health information (17–20) Evidence from other settings suggests that culturally tailored health education can improve knowledge and engagement compared with standard approaches (21). This may be particularly relevant where biomedical concepts have no direct linguistic or cultural equivalents or where cultural understandings influence how pregnancy, childbirth, and danger signs are understood (18).

The Hehe population in the Iringa Region provides an important context for culturally and linguistically appropriate BPCR education. Although Swahili is widely used in health services, Kihehe remains an important linguistic and cultural reference point in many rural communities, including those in Kilolo and Mufindi districts (22). Therefore, the expression and explanation of BPCR information may influence women’s understanding of danger signs and other essential BPCR concepts.

Despite the widespread provision of BPCR education through antenatal care, evidence remains limited on whether culturally adapted educational materials improve BPCR knowledge compared with standard translated materials in rural Tanzania. Few studies have directly compared a Culturally Adapted Swahili BPCR Training Manual with a Standard Translated Swahili BPCR Training Manual while maintaining comparable core educational content (23).

Therefore, this study aimed to evaluate the effect of a previously prepared Culturally Adapted Swahili BPCR Training Manual, compared with a Standard Translated Swahili BPCR Training Manual, on BPCR knowledge among Hehe pregnant women attending antenatal care in the rural Iringa Region, Tanzania.

## Methods

### Study design and setting

A controlled pretest–posttest quasi-experimental study was conducted to evaluate two previously prepared BPCR educational packages. The intervention group received the Culturally Adapted Swahili BPCR Training Manual, whereas the control group received the Standard Translated Swahili BPCR Training Manual.

The study was guided by the Health Belief Model. The educational intervention emphasized understanding pregnancy and childbirth complications, recognizing danger signs, the importance of advance preparation, the benefits of timely care-seeking, and actions to overcome barriers to obtaining appropriate maternal healthcare (24).

The study was conducted in Kilolo and Mufindi districts of Iringa Region, southern Tanzania. The area is predominantly rural and largely inhabited by the Hehe population. Maternal healthcare services are provided through dispensaries, health centers, and hospitals, with antenatal care serving as an important entry point to the health system (25). Baseline data were collected from 18 to 21 November 2025, and post-intervention data were collected from 16 to 19 December 2025. One district was designated as the intervention district and the other as the control district. The geographical separation of the study arms was intended to reduce information exchange and contamination between participants receiving the different educational packages.

### Participants

Eligible participants were pregnant women attending antenatal care at selected public health facilities who were in the first or second trimester, belonged to the Hehe population, had resided in the Iringa Region for at least six months, were willing to participate, and provided informed consent.

Women in the third trimester were excluded because the study required sufficient time to participate in the educational intervention and complete the subsequent follow-up assessment. Women employed in the health sector were excluded because of their potential prior exposure to maternal health education. Women enrolled in other research or health programs that could interfere with the intervention were also excluded. Participants who developed serious medical or obstetric conditions that prevented continued participation during the study period were excluded from further participation.

### Sample size and sampling

The sample size was estimated using WinPEPI version 11.65, developed by Abramson in 2011, based on a comparison of mean BPCR knowledge scores between two independent groups, using a two-sided significance level of 0.05 and 80% statistical power (26). The expected difference in BPCR knowledge scores and estimates of outcome variability were informed by a previous quasi-experimental BPCR intervention study conducted in Indonesia by Astika et al. (27).

The previous study reported a mean BPCR knowledge score of d 83.1 (SD 5.2) in the intervention group and 55.1 (SD 3.4) in the control group. These estimates were used as the expected outcome means and variability parameters in the WinPEPI calculation. The calculation produced a minimum required sample size of 164 participants. A total of 176 pregnant women were recruited, comprising 88 participants in the intervention group and 88 in the control group.

### Sampling technique and procedure

A multistage sampling procedure was used to select study sites and participants. The sampling process involved five stages: selection of districts, divisions, wards, villages, and participants.

Three rural districts in the Iringa Region were eligible for inclusion, from which two districts were randomly selected using simple random sampling. The two selected districts were Mufindi and Kilolo. Mufindi was assigned to the intervention group, while Kilolo was assigned to the control group. The intervention district received the Culturally Adapted Swahili BPCR Training Manual, whereas the control district received the Standard Translated Swahili BPCR Training Manual. Within each selected district, divisions, wards, and villages were selected using simple random sampling. Eligible pregnant women were then identified from antenatal care registers and selected using systematic random sampling.

#### Stage 1: Selection of districts

Three rural districts in the Iringa Region were eligible for inclusion. Two districts, Mufindi and Kilolo, were randomly selected using simple random sampling. Mufindi was assigned to the intervention group, while Kilolo was assigned to the comparison group. The intervention district received the Culturally Adapted Swahili BPCR Training Manual, whereas the comparison district received the Standard Translated Swahili BPCR Training Manual. Thus, each study condition was represented by one district.

#### Stage 2: Selection of divisions

Each selected district had four divisions. Two divisions were selected from each district using simple random sampling through a lottery method, resulting in four selected divisions. An independent person conducted the random selection. The selected divisions formed the sampling units for subsequent ward selection.

#### Stage 3: Selection of wards

Two wards were selected from each of the four selected divisions using simple random sampling through a lottery method, resulting in eight selected wards. An independent person conducted the random selection of wards from each selected division.

#### Stage 4: Selection of villages

Two villages were selected from each of the eight selected wards using simple random sampling through a lottery method, resulting in 16 selected villages. An independent person conducted the random selection. Eight villages were therefore selected from the intervention district and eight from the comparison district. The 16 selected villages constituted the geographical sampling units for participant recruitment.

#### Stage 5: Selection of participants

Public dispensaries providing antenatal care services to women from the selected villages were identified as participant recruitment sites, and all eligible dispensaries serving the selected villages were included. Pregnant women attending antenatal care at the selected dispensaries were assessed for eligibility using the predefined inclusion and exclusion criteria. Eligibility, including self-identification as Hehe, was confirmed with each woman before recruitment.

A total of 176 participants were recruited, with 11 participants selected from each of the 16 selected villages. Thus, eight intervention villages contributed 88 participants, and eight comparison villages contributed 88 participants. Eligible women were selected using systematic random sampling. For each village, the sampling interval (k) was calculated as k = N/n, where N represented the total number of eligible pregnant women in the sampling frame and n represented the target sample of 11 participants. A random starting participant was selected, followed by every kth eligible woman until 11 participants were recruited from each village. The same procedure was applied in both intervention and comparison villages.

### Data collection procedures

Baseline data were collected from 18 to 21 November 2025 before delivery of the educational intervention. Trained research assistants who were experienced midwives conducted face-to-face interviews in private and quiet areas of the participating antenatal care clinics. Each interview lasted approximately 15–20 minutes. Research assistants received standardized training on questionnaire administration, ethical procedures, interview techniques, confidentiality, and neutrality. Personnel responsible for intervention delivery were separate from those responsible for outcome assessment. Participants were assigned unique identification numbers, and names were not recorded on the study questionnaires.

Post-intervention data were collected from 16 to 19 December 2025, four weeks after baseline assessment, using the same questionnaire and data collection procedures.

### Data collection instrument

A structured questionnaire adapted from standardized BPCR monitoring tools developed by the Johns Hopkins Program for International Education in Gynecology and Obstetrics (JHPIEGO) was used (10). The questionnaire collected sociodemographic and obstetric characteristics and assessed BPCR knowledge across key domains, including birth preparedness, danger signs during pregnancy, labor and childbirth, the postnatal period, and newborn danger signs.

### BPCR knowledge

BPCR knowledge was assessed using 64 dichotomous items covering danger signs during pregnancy, danger signs during labor and childbirth, danger signs during the postnatal period, newborn danger signs, birth preparedness, and antenatal care. Each correct response was scored 1, and each incorrect response was scored 0. The total knowledge score ranged from 0 to 64, with higher scores indicating greater BPCR knowledge.

### Validity and reliability

The questionnaire was adapted from standardized BPCR assessment tools previously used in maternal health research and monitoring surveys among women in low- and lower-middle-income countries, including Tanzania (10). Similar BPCR assessment tools have also been used in Malaysia, where the attitude scale demonstrated a Cronbach’s alpha of 0.72 (28), and in Ethiopia, where a Cronbach’s alpha of 0.85 was reported (29).

The questionnaire underwent translation, expert review, and pretesting before use in the study. The English questionnaire was translated into Swahili by bilingual experts. Independent back-translation was subsequently undertaken, and discrepancies between the original and translated versions were reviewed and resolved by the research team and language experts.

Pretesting was conducted among pregnant women outside the study area to assess the clarity, comprehensibility, and appropriateness of the questionnaire. Internal consistency was assessed using data collected in the present study. Because the knowledge items were dichotomously scored, reliability was assessed using the Kuder–Richardson Formula 20 (KR-20). The overall knowledge scale demonstrated strong internal consistency (KR-20 = 0.892).

### Study variables

The primary independent variable was study group assignment, categorized as the intervention group receiving the Culturally Adapted Swahili BPCR Training Manual and the control group receiving the Standard Translated Swahili BPCR Training Manual. Participant characteristics considered as covariates included maternal age, gestational age, gravidity, marital status, living arrangement, educational level, reading ability, and occupation. The dependent variable was BPCR knowledge, operationalized as the total score ranging from 0 to 64. Higher scores indicated greater BPCR knowledge.

### Implementation of the educational packages

The two educational packages evaluated in this study had been prepared before the present evaluation. The present study focused on their implementation and comparative effect on BPCR knowledge. The intervention and control conditions were standardized with respect to the number and duration of sessions, delivery schedule, teaching format, language of instruction, and follow-up procedures. The principal difference between the two study conditions was the educational package used.

### Culturally adapted Swahili BPCR training manual

After completion of the baseline assessment, participants in the intervention group received the Culturally Adapted Swahili BPCR Training Manual. Each session was facilitated by one trained intervention trainer using the training manual, timetable, and corresponding Swahili lesson plan. Each session lasted approximately two hours, comprising approximately one hour of structured facilitation and one hour of interactive discussion, questions, and answers, as specified in the implementation timetable.

The sessions were delivered face-to-face in Swahili using a participatory approach. The trainer presented BPCR information using the culturally adapted terminology, contextual explanations, and visual representations incorporated into the training manual. For concepts without culturally equivalent terminology, visual aids were used together with explanatory descriptions to facilitate participants’ understanding. Participants were encouraged to ask questions and discuss the session content, and the trainer provided clarification in Swahili to support comprehension and active participation.

### Standard translated Swahili BPCR training manual

Participants in the control group received the previously prepared Standard Translated Swahili BPCR Training Manual. The control sessions were delivered using the same number of sessions, session duration, schedule, face-to-face format, participatory teaching approach, and Swahili language of instruction as those used in the intervention group.

The sessions covered the same core BPCR subject areas, including birth preparedness, danger signs, skilled birth attendance, emergency preparedness, antenatal care, and timely healthcare-seeking. The principal difference between the two conditions was the educational package: the intervention group received the Culturally Adapted Swahili BPCR Training Manual, whereas the control group received the Standard Translated Swahili BPCR Training Manual.

At the end of the educational sessions, participants in both groups were informed of the scheduled four-week follow-up assessment, which coincided with their next scheduled monthly antenatal care visit. The follow-up date was recorded in their clinic cards as a reminder. After completing the educational sessions, participants received their routine antenatal care services from a midwife at the health facility.

### Evaluation phase

Four weeks after implementation of the assigned educational package, post-intervention assessment was conducted among participants in both groups from 16 to 19 December 2025. The same structured questionnaire and face-to-face interview procedures used at baseline were applied at the post-intervention assessment. Different research assistants conducted the post-intervention assessment from those who conducted the baseline assessment. Assessments were conducted at the participating antenatal care facilities in private and quiet areas. The same procedures for questionnaire administration, confidentiality, and participant identification used at baseline were maintained during the post-intervention assessment.

### Intervention fidelity and contamination control

The intervention and control sessions were delivered separately at different locations on the same dates. The number and duration of sessions, delivery schedule, teaching format, language of instruction, and follow-up procedures were standardized across the two study groups. Trainers assigned to the intervention group did not participate in delivery of the control educational sessions. Data collectors were separate from the trainers and were not involved in delivery of either educational package. Different research assistants conducted the baseline and post-intervention assessments.

The researcher provided overall supervision and administrative oversight of the study, including distribution of questionnaires and provision of financial, stationery, and technical support, but did not administer the questionnaires, deliver either educational package, or participate in baseline or post-intervention data collection.

To minimize contamination, the intervention and control groups were implemented in geographically separate districts. Outcome assessment was conducted by research assistants who were not involved in the delivery of either educational package. The same questionnaire, assessment schedule, and researcher-administered questionnaire procedures were used in both groups.

### Data analysis

Data were analyzed using IBM Statistical Package for the Social Sciences version 27. Descriptive statistics were used to summarize participant characteristics and BPCR knowledge scores. Continuous variables were summarized using means and standard deviations, while categorical variables were summarized using frequencies and percentages. The distribution of the continuous outcome was assessed before inferential analysis, and parametric methods were used when the relevant assumptions were considered acceptable.

Baseline characteristics of the intervention and control groups were compared using independent-samples t-tests for continuous variables and chi-square tests for categorical variables. These comparisons were used to describe baseline characteristics and assess differences between the study groups before the intervention.

### Difference-in-differences analysis

The primary analysis used a Difference-in-Differences regression approach to estimate whether the change in BPCR knowledge from baseline to the four-week post-intervention assessment differed between the intervention and control groups. The study group was coded as 0 for the control group and 1 for the intervention group, while time was coded as 0 for baseline and 1 for post-intervention. The model included the study group, time, and the Group × Time interaction:

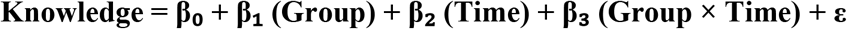

The Group × Time interaction coefficient (β₃) represented the DiD estimate and therefore quantified the difference in change in BPCR knowledge between the intervention and control groups. Standard errors and 95% confidence intervals were estimated using the regression procedure applied in the analysis.

### Adjusted analysis

Because BPCR knowledge was analyzed as a continuous outcome, multiple linear regression was used to examine whether intervention-group assignment was associated with post-intervention knowledge after adjustment for baseline knowledge and selected participant characteristics. The model included study group, baseline knowledge score, maternal age, educational level, gravidity, marital status, occupation, living arrangement, and reading ability. Adjusted regression coefficients were reported with 95% confidence intervals and p-values.

Statistical significance was set at p < 0.05. No missing outcome data were identified; therefore, no imputation procedures were required. No post-hoc subgroup or sensitivity analyses were conducted.

### Ethical clearance and informed consent

As the study involved human participants, ethical clearance was obtained from the Research Review Committee of the Research and Publication Office at the University of Dodoma prior to data collection (reference number MA.84/261/86/11). All procedures involving human participants were conducted in accordance with institutional ethical standards and the principles of the Declaration of Helsinki.

Permission to conduct the study in the Iringa Region was obtained from the Permanent Secretary, President’s Office, Regional Administration and Local Government (reference number AB.307/323/01/R/70). Permission to collect data in the Iringa Region was also obtained from the Iringa Regional Secretary (reference number Na.FA.225/265/01/J/281). Permission to conduct the study in Mufindi and Kilolo districts was obtained from the respective district directors. The permission reference number for Mufindi District was Na. HW/MUF/S.40/43/VOL.IV/39, and that for Kilolo District was Na. KDC/S.10/4/VOLL.VIII/193. Permission to use the antenatal care guidelines for the study was obtained from the Ministry of Health (reference number PA.104/282/02).

Written informed consent was obtained from all participants before inclusion in the study. Participants were informed about the purpose and procedures of the study, potential benefits, and the voluntary nature of participation. They were informed that they could withdraw from the study at any time without affecting their access to healthcare services. Confidentiality and privacy were maintained throughout the study by assigning unique identification numbers instead of recording participants’ personal names.

Cultural respect and sensitivity were observed throughout all stages of the study to ensure that the training content, delivery, and implementation were culturally appropriate and did not conflict with participants’ beliefs, values, or customary practices.

## Results

A total of 176 pregnant women were enrolled, with 88 participants allocated to the Culturally Adapted BPCR Training Manual intervention group and 88 allocated to the Standard Translated BPCR Training Manual control group. All enrolled participants received the assigned educational program and completed both baseline and post-intervention assessments. There were no losses to follow-up or exclusions after allocation, and all 176 participants were included in the final analysis. No adverse events or unintended consequences related to the educational interventions were reported in either group.

### Sociodemographic characteristics of study participants

Table 1 presents the baseline sociodemographic and obstetric characteristics of the 176 study participants by study group. Most participants were aged 20–34 years, were between 13 and 24 weeks of gestation, were multigravida, were married, and were living with a partner. The majority had primary education, reported reading easily, and were engaged in farming as their main occupation (Table 1).

**Table 1:** Baseline sociodemographic characteristics of pregnant women by study group (N = 176).

| Variables | Category | Intervention (n = 88)<br>n (%) | Control (n= 88)<br>n (%) |
| --- | --- | --- | --- |
| **Age (years) Mean±SD (Min-Max)** |  | 26.50 ± 6.35 (18-44) | 26.38 ± 5.89 (18-42) |
|  | <20 | 15 (17.0) | 13 (14.8) |
|  | 20-34 | 60 (68.2) | 65 (73.9) |
|  | ≥35 | 13 (14.8) | 10 (11.4) |
| **Gestational age (months) Mean ± SD (Min-Max)** |  | 4.80 ± 1.13 (2-6) | 4.53 ± 1.29 (2-6) |
|  | ≤12 weeks | 11 (12.5) | 20 (22.7) |
|  | 13-24 weeks | 77 (87.5) | 68 (77.3) |
| **Gravidity** | Primigravida | 21 (23.9) | 10 (11.4) |
|  | Multigravida | 67 (76.1) | 78 (88.6) |
| **Marital status** | Single | 15 (17.0) | 8 (9.1) |
|  | Married | 73 (83.0) | 80 (90.9) |
| **Living with a partner** | No | 18 (20.5) | 10 (11.4) |
|  | Yes | 70 (79.5) | 78 (88.6) |
| **Educational level** |  |  |  |
|  | Primary education | 57 (64.8) | 64 (72.7) |
|  | Secondary education and above | 31 (35.2) | 24 (27.3) |
| Reading ability | Easily | 82 (93.2) | 86 (97.7) |
|  | With difficulty | 6 (6.8) | 2 (2.3) |
| Occupation | Peasant | 78 (88.6) | 80 (90.9) |
|  | Business woman/employed | 10 (11.4) | 8 (9.1) |

### Baseline comparison of knowledge between intervention and control groups

At baseline, the mean BPCR knowledge score was 21.01 ± 7.42 in the intervention group and 22.51 ± 5.80 in the control group, with no statistically significant difference between the groups (mean difference = −1.50, p = 0.137).

### Post-intervention comparison of BPCR knowledge between intervention and control groups

At post-intervention, the intervention group had significantly higher mean BPCR knowledge scores than the control group (38.49 ± 7.54 vs. 32.70 ± 5.61; mean difference = 5.79, p < 0.001).

### Within-group comparison of birth BPCR knowledge before and after the intervention

Within-group comparisons showed significant improvements in knowledge in the intervention group following the intervention. Knowledge scores increased from 21.01 ± 7.42 at pretest to 38.49 ± 7.54 at the posttest, representing a mean increase of 17.48 points (t = 14.577, p < 0.001) (Table 2).

**Table 2:** Within-group comparison of BPCR knowledge before and after the intervention among pregnant women (N = 176).

| Group | Posttest Mean ± SD | Pretest Mean ± SD | Mean difference (posttest – pretest) | t | P-Value |
| --- | --- | --- | --- | --- | --- |
| **Knowledge** |  |  |  |  |  |
| Intervention | $38.49 \pm 7.54$ | $21.01 \pm 7.42$ | 17.48 | 14.577 | <0.001 |
| Control | $32.70 \pm 5.61$ | $22.51 \pm 5.80$ | 10.19 | 12.994 | <0.001 |

### Factors associated with post-test knowledge scores

In the adjusted multiple linear regression analysis, assignment to the intervention group was significantly associated with higher post-test knowledge scores (β = 5.880, 95% CI: 3.841–7.920, *p* < 0.001). After adjustment for education, baseline knowledge, gravidity, marital status, living with a partner, reading ability, occupation, and age, participants in the intervention group had approximately 5.88 points higher post-test knowledge scores than those in the control group. None of the other variables included in the model were significantly associated with post-test knowledge scores Table 3.

**Table 3:**
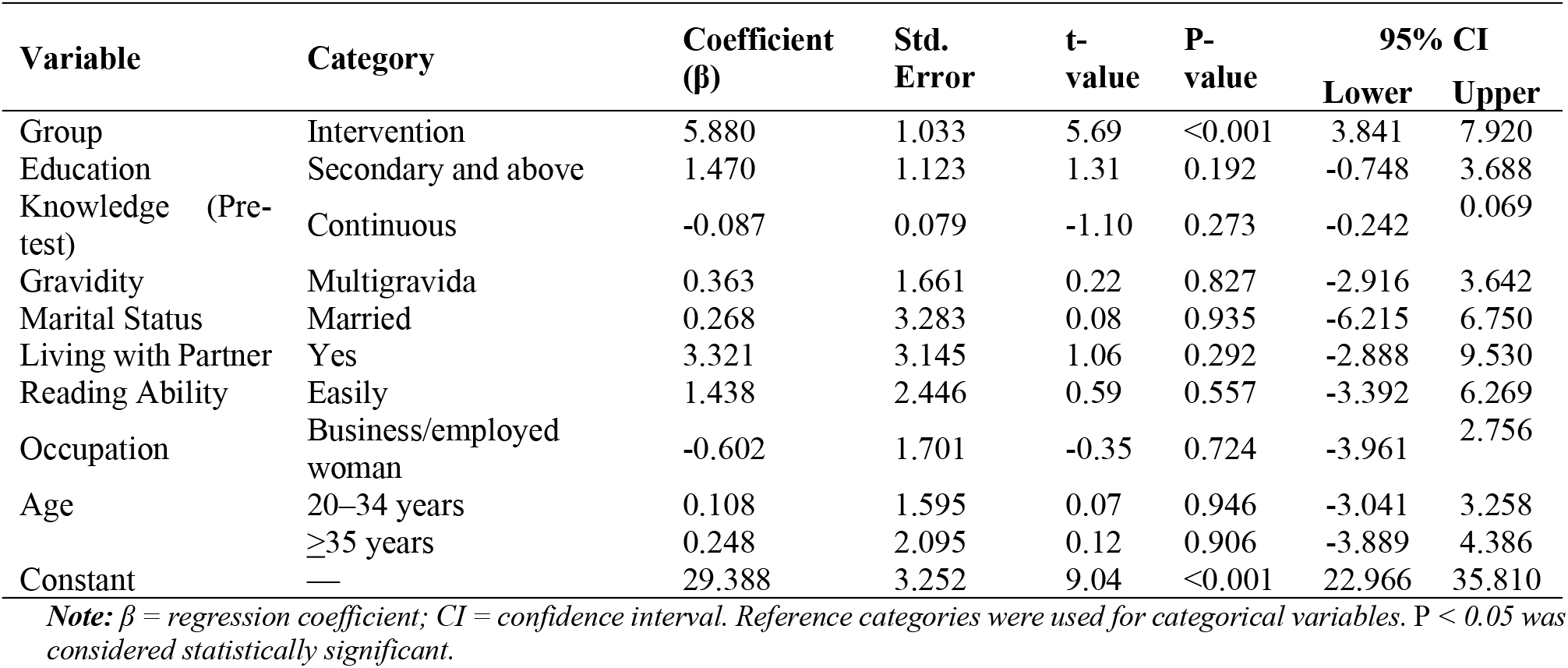
Adjusted multiple linear regression analysis of factors associated with post-test knowledge scores among pregnant women (N = 176)

### Difference-in-Differences regression analysis of the effect of culturally adapted BPCR training on knowledge

The Difference-in-Differences regression analysis showed that the culturally adapted BPCR training was associated with a significantly greater improvement in BPCR knowledge in the intervention group compared with the control group. The Group × Time interaction was positive and statistically significant (β = 7.18, 95% CI: 4.40–9.97, p < 0.001), indicating that the increase in BPCR knowledge from baseline to post-intervention was approximately 7.18 points greater in the intervention group than in the control group Table 4.

**Table 4:**
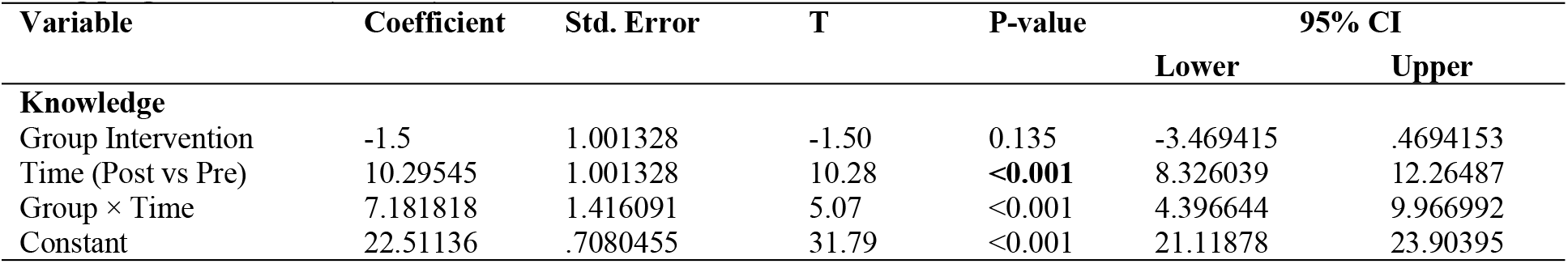
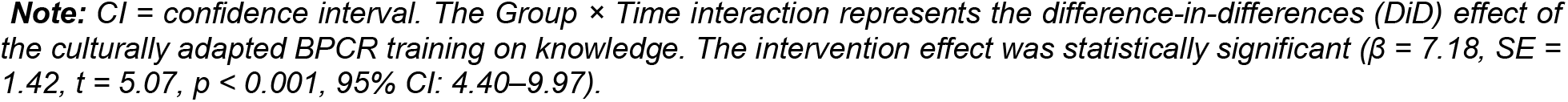
Difference-in-Differences regression analysis of the effect of culturally adapted BPCR training on knowledge among pregnant women (N = 176)

| Variable | Coefficient | Std. Error | T | P-value | 95% CI |  |
| --- | --- | --- | --- | --- | --- | --- |
|  |  |  |  |  | Lower | Upper |
| **Knowledge** |  |  |  |  |  |  |
| Group Intervention | -1.5 | 1.001328 | -1.50 | 0.135 | -3.469415 | .4694153 |
| Time (Post vs Pre) | 10.29545 | 1.001328 | 10.28 | <0.001 | 8.326039 | 12.26487 |
| Group × Time | 7.181818 | 1.416091 | 5.07 | <0.001 | 4.396644 | 9.966992 |
| Constant | 22.51136 | .7080455 | 31.79 | <0.001 | 21.11878 | 23.90395 |

**Note:** CI = confidence interval. The Group × Time interaction represents the difference-in-differences (DiD) effect of the culturally adapted BPCR training on knowledge. The intervention effect was statistically significant ( $\beta = 7.18$ , $SE = 1.42$ , $t = 5.07$ , $p < 0.001$ , 95% CI: 4.40–9.97).

### Estimated marginal means of knowledge by group and time

The estimated marginal means showed an increase in BPCR knowledge from pre- to post-intervention in both groups, with a greater increase in the intervention group. Knowledge increased from 21.01 to 38.49 in the intervention group, compared with an increase from 22.51 to 32.81 in the control group Table 5.

**Table 5:** Estimated marginal means of BPCR knowledge by group and time among pregnant women (N = 176).

| Knowledge: |  | Margin | Std. Error | t-value | P-value | 95% CI |  |
| --- | --- | --- | --- | --- | --- | --- | --- |
| Group | Time |  |  |  |  | Lower | Upper |
| Control | Pre | 22.51136 | .7080455 | 31.79 | <0.001 | 21.11878 | 23.90395 |
| Control | Post | 32.80682 | .7080455 | 46.33 | <0.001 | 31.41423 | 34.19941 |
| Intervention | Pre | 21.01136 | .7080455 | 29.68 | <0.001 | 19.61878 | 22.40395 |
| Intervention | Post | 38.48864 | .7080455 | 54.36 | <0.001 | 37.09605 | 39.88122 |

## Discussion

This study found that culturally adapted BPCR education was associated with a greater improvement in BPCR knowledge than standard translated BPCR education among Hehe pregnant women attending antenatal care in rural Iringa. Although knowledge increased in both groups, the improvement was substantially greater in the intervention group. The Difference-in-Differences analysis indicated that the intervention group experienced a 7.18-point greater increase in knowledge than the control group (95% CI: 4.40–9.97; *p* < 0.001), and this association remained statistically significant after adjustment for baseline knowledge and participant characteristics. These findings suggest that culturally adapted BPCR education may provide an additional benefit for improving knowledge beyond standard translated education.

The increase in BPCR knowledge in both groups is consistent with evidence that structured maternal health education can improve women’s knowledge of pregnancy, childbirth, postpartum, and newborn danger signs (19, 30). The results were similar, with a quasi-experimental study in rural Tanzania evaluating the education BPCR intervention, which reported significant improvements in knowledge of maternal danger signs and a positive net intervention effect compared with a comparison district (31). Similarly, studies from other sub-Saharan African settings have reported improvements in knowledge of obstetric danger signs and birth preparedness following structured or context-specific maternal health education (19). These findings support the role of structured BPCR education within antenatal care in addressing knowledge gaps related to the recognition of maternal and newborn complications.

Importantly, the present study extends this evidence by examining cultural adaptation in addition to education itself. Both study groups received BPCR education under standardized conditions, including the same language, number and duration of sessions, delivery schedule, teaching format, and follow-up procedures. The principal difference between the groups was the educational package used: the intervention group received the Culturally Adapted Swahili BPCR Training Manual, whereas the control group received the Standard Translated BPCR Training Manual. Therefore, the greater improvement observed in the intervention group may be related to the cultural and linguistic adaptation of BPCR concepts rather than simply to greater exposure to education. Culturally familiar terminology, contextual explanations, and locally meaningful examples may have made some BPCR concepts easier for participants to understand and recall at the post-intervention assessment. Previous research has suggested that culturally and linguistically adapted health education can improve comprehension and health knowledge by making health information more meaningful and accessible to the intended population (32, 33). A systematic review of culturally appropriate maternal health education in low- and middle-income countries similarly reported beneficial effects of contextually adapted interventions on maternal knowledge and related outcomes, although substantial heterogeneity in intervention content and methodological quality was reported (34, 35).

The relatively low baseline knowledge observed in both groups provides further context for the observed improvement. The baseline mean knowledge scores were 21.01 out of 64 in the intervention group and 22.51 out of 64 in the control group, indicating that BPCR knowledge was relatively low before the educational intervention. This pattern is consistent with previous evidence from rural Iringa showing limited BPCR knowledge among pregnant women, including important gaps in the recognition of postpartum, newborn, and labor-related danger signs (9). Similar knowledge gaps have been reported among pregnant women in Tanzania and other sub-Saharan African settings (9, 36, 37). Limited recognition of maternal and newborn danger signs is an important educational concern because knowledge of potential complications is relevant to recognizing when further assessment or care may be required (38, 39). The improvement observed following BPCR education therefore represents progress in addressing an identified knowledge gap in this population.

The significant improvement in the control group also warrants consideration. Knowledge increased by approximately 10.29 points among women who received the Standard Translated BPCR Training Manual. This indicates that standard BPCR education was also associated with an improvement in knowledge and should not be interpreted as an ineffective educational approach. Rather, the significant Group × Time interaction indicates that the improvement in the intervention group was greater than the improvement observed with the standard translated package. This distinction is important because the study compared two educational approaches containing comparable core BPCR content rather than comparing education with no education. The findings therefore suggest that, in addition to the content of maternal health education, the way information is linguistically and culturally expressed may be associated with the magnitude of knowledge gained (30).

### Theoretical implications

The findings have implications for the Health Belief Model, which provided a framework for interpreting how knowledge may relate to maternal health decision-making. The Health Belief Model proposes that health actions are influenced by perceived susceptibility, severity, benefits, barriers, cues to action, and self-efficacy (40). Although these constructs were not directly measured, improved BPCR knowledge may strengthen women’s understanding of maternal and newborn complications and the importance of preparation and timely response.

Culturally adapted communication may facilitate understanding of Health Belief Model-related concepts (40). Women may be able to spot problems more easily if they know what danger signs to look for. On the other hand, explanations of transportation, finances, skilled birth attendance, and emergency plans may help them understand the benefits of being ready for birth (41). Contextual explanations may also reduce informational barriers to recommended actions (34, 41). Thus, the findings support incorporating cultural and linguistic adaptation into the Health Belief Model-informed approach.

However, improved knowledge does not demonstrate changes in Health Belief Model constructs or subsequent behavior. This study did not measure perceived susceptibility, severity, benefits, barriers, cues to action, or self-efficacy (40). Future studies should assess these constructs and examine whether culturally adapted BPCR education influences perceptions, preparedness, and care-seeking behaviors (43).

### Practical implications

The findings suggest that BPCR education may benefit from approaches extending beyond direct translation. Cultural and linguistic adaptation can express biomedical concepts using locally understandable terminology and examples, particularly for danger signs and concepts lacking familiar linguistic equivalents (44). Culturally adapted BPCR materials could be incorporated into routine ANC group education, individual counselling, community maternal-health programmes, and provider training. Adaptation should preserve biomedical meaning while using understandable terminology, contextual explanations, and appropriate visual materials (45). Healthcare providers may also benefit from training in culturally responsive communication to explain difficult concepts without compromising clinical accuracy (46). At the program level, health authorities could integrate cultural adaptation into maternal-health education material development through collaboration among health professionals, language experts, community representatives, and target populations, supporting technical accuracy and comprehensibility (47).

### Strengths of the study

The study compared two active educational approaches rather than the culturally adapted intervention with no education, allowing assessment of additional improvement beyond the standard translated package. The groups had comparable session number and duration, delivery schedule, teaching format, language, and follow-up, making the educational package the principal planned difference. Complete follow-up was achieved, with all 176 participants completing baseline and post-intervention assessments, reducing potential attrition bias. The knowledge instrument demonstrated strong internal consistency (KR-20 = 0.892). Difference-in-Differences regression compared changes over time between groups rather than relying only on post-intervention differences (47). Consistent findings across post-intervention comparisons, within-group analyses, adjusted regression, and Difference-in-Differences regression analysis further support the observed association between the culturally adapted package and improved knowledge.

### Limitations of the study

First, the quasi-experimental design did not involve individual randomization. Although districts were selected using random sampling, allocation to study conditions was not equivalent to individual randomization; residual confounding cannot be completely excluded despite adjustment for measured characteristics. Second, Difference-in-Differences regression analysis used only one pre-intervention measurement, preventing empirical assessment of the parallel-trends assumption (48). Although baseline knowledge did not differ statistically between groups, this does not establish complete baseline equivalence or exclude unmeasured differences. DiD findings were therefore interpreted alongside adjusted regression and other analyses (49).

## Conclusion

Culturally adapted BPCR education was associated with a greater increase in BPCR knowledge than standard translated education among Hehe pregnant women in rural Iringa. However, the quasi-experimental design, one district per study arm, and single pre-intervention measurement limit causal inference. The findings support consideration of cultural and linguistic adaptation in BPCR educational materials for similar populations. The study assessed knowledge but not Health Belief Model constructs, preparedness, or care-seeking behavior. Future research should use multisite designs with more than one district per arm, stronger allocation procedures, multiple baseline measurements, and longer follow-up to assess the durability and transferability of knowledge gains and examine associations with Health Belief Model constructs and maternal health behaviors.

## Data Availability

The datasets generated and analyzed during the current study are not publicly available because they contain information that could potentially identify participants.

## Trial registration

Pan African Clinical Trials Registry PACTR202607626025411, registered retrospectively on 09 July 2026 https://pactr.samrc.ac.za

## Consent for publication

Not applicable.

## Availability of data and materials

The datasets generated and analyzed during the current study are not publicly available because they contain information that could potentially identify participants. However, de-identified data may be available from the corresponding author upon reasonable request for legitimate academic or research purposes, subject to approval by the relevant ethics committee, institutional policies, and applicable participant consent provisions.

## Acknowledgments

The authors sincerely thank the Ministry of Health, Tanzania, for granting permission to pursue PhD studies at the University of Dodoma. We also gratefully acknowledge the University of Dodoma for providing ethical approval and institutional support that facilitated the conduct and dissemination of this research. We extend our sincere appreciation to all study participants for their time, willingness to participate, and valuable contributions to the study. We also thank the health-care providers and local authorities in the Iringa Region for their cooperation and support during the study.

## Authors’ contributions

RBM contributed to conceiving the study and conceptualization, writing the background information, study design, approaches and study methodology, data collection, software data analysis, data curation and visualization, project supervisor and funding, and writing the original draft and refined version. FVM, SAS, and RS contributed to the background, study design, approaches, and methodology, as well as data analysis and visualization; the manuscript was refined with input from all authors.

